# Asthma-like bronchodilator responsiveness in patients with neuronal intranuclear inclusion disease

**DOI:** 10.64898/2025.12.11.25342120

**Authors:** Daisuke Tahara, Nao Tahara, Chisato Tamai, Akio Akagi, Yuichi Riku, Hiroaki Miyahara, Rei Kobayashi, Hisashi Okada, Michi Kawamoto, Junko Ishii, Hiroki Yamazaki, Takashi Kurashige, Atsuhiko Sugiyama, Akiko Nagaishi, Katsuya Nishida, Kazuma Sugie, Takayasu Fukudome, Kazuko Hasegawa, Hiroyuki Ishiura, Haruki Koike, Takashi Kasai, Toshiki Mizuno, Masahiro Ando, Yujiro Higuchi, Fumiaki Tanaka, Yuishin Izumi, Gen Sobue, Yasushi Iwasaki, Satoru Ito, Jun Sone, the Japanese Consortium for neuronal intranuclear inclusion disease study (JaNIIIDS) group

## Abstract

Neuronal intranuclear inclusion disease (NIID) is a neurodegenerative condition characterized by the presence of intranuclear inclusions in neuronal and visceral cells. Patients with NIID can present respiratory symptoms; however, data on pulmonary functions in NIID are lacking. This study investigated the respiratory conditions in NIID patients diagnosed with histopathological and genetic studies. We conducted two spirometries before and after administrating the bronchodilator in NIID patients with asthmatic histories or symptoms. We statistically compared pre- and post-measured values including forced vital capacity (FVC), forced expiratory volume in one second (FEV_1_), and peak expiratory flow (PEF). Before two spirometries, we also measured fractional concentration of exhaled nitric oxide (FeNO), if possible. Of the 51finally enrolled patients, 17 (33.3%, 95% CI 20.8% to 47.9%) patients had asthmatic histories or symptoms, and 14 patients received two spirometries. After administrating the bronchodilator, FEV_1_ and PEF significantly increased by 150 mL (6.01%, *p* = 0.002) and 260 mL/s (6.72%, *p* = 0.017), respectively. The median (interquartile range) of FeNO measured in nine patients was 15 (10–21) ppb. Patients with NIID have airflow reversibility like asthma. Airway inflammation is less associated with this condition; thus, immunomodulators such as corticosteroid may not improve respiratory symptoms in NIID.

## Introduction

Neuronal intranuclear inclusion disease (NIID) is a neurodegenerative disorder which is characterized by the presence of eosinophilic intranuclear inclusion in central and peripheral neurons, glial cells, and visceral cells [1, 2]. This condition is also called as “neuronal intranuclear hyaline inclusion disease” [1] or “intranuclear inclusion body disease” [3]. The antemortem diagnosis of NIID had been difficult; however, the skin biopsy was confirmed useful for diagnosing NIID [4], and later GGC repeat expansions in the *NOTCH2NLC* gene were identified in patients with NIID [5, 6]. Due to the improvements of diagnosis methods, the number of patients antemortem diagnosed as NIID has been recently increasing. The prevalence rate of NIID is estimated higher than previously thought [2], so nowadays revealing its clinical and pathological characteristics has been become increasingly important.

Patients with NIID present various nervous symptoms including dementia, muscle weakness, sensory disturbance, encephalic episodes, and retinopathy [2, 7]. It is thought that autonomic symptoms (e.g. miosis and bladder dysfunction) occur because the sympathetic system is dominantly disturbed in NIID patients [2, 7, 8]. In addition to nervous symptoms, other systemic symptoms such as IgA nephropathy [9] and cardiomyopathy [10] can be observed.

In our NIID cohort, we found many NIID patients who had been diagnosed with bronchial asthma at other hospitals. Furthermore, many NIID patients had a history of receiving treatment for asthma but discontinued it because they felt it was not effective. We became interested in the relationship between NIID and asthma and began to investigate the relationship. According to a paper from China, respiratory symptoms are the most common initial symptoms followed by nervous symptoms in patients with GGC repeat expansions in the *NOTCH2NLC* gene [11]. Some studies reported patients with NIID having past medical histories of asthma [12, 13], and another study showed three out of four patients with NIID had chronic coughing [14]. It seems informative to obtain the respiratory conditions in patients with NIID; however, comprehensive data are lacking.

Spirometry is one of the physiological tests for evaluating objectively respiratory conditions. By the slow manoeuvre, vital capacity (VC) and inspiratory capacity (IC) can be examined [15]. By the forced manoeuvre, forced vital capacity (FVC), forced expiratory volume in one second (FEV_1_), peak expiratory flow (PEF), and maximal mild-expiratory flow (MMF) are examined [15]. FVC is the total forced expiratory volume from full inspiration, and the expiratory volume in the first second of this manoeuvre is FEV_1_ [15]. PEF is the maximal expiratory flow in a FVC manoeuvre [15], and MMF is the expiratory flow between 25% and 75% of the FVC [16]. Generally, FEV_1_/FVC < 70% is interpreted as the presence of obstructive ventilatory disorders, such as asthma and chronic obstructive pulmonary disease (COPD) [17]. Additionally, bronchodilator responsiveness (BDR) testing is useful for the diagnosis of obstructive ventilatory disorders [15, 17]. In BDR testing, two spirometries are conducted before and after the administration of the bronchodilator [15].

The purpose of the current study is to investigate the respiratory conditions and pathology in NIID patients by means of history taking, spirometry, and BDR testing.

## Methods

### Study design and participants

This multicentre cross-sectional study in Japan included patients with NIID who had previously participated in research of the Japanese Consortium for neuronal intranuclear inclusion disease study (JaNIIDS). NIID patients included in this study were diagnosed by histopathological studies including ante-mortem skin biopsy [4] and other histopathological specimen such as surgically removed specimens, and genetic test confirming GGC repeat expansion in the *NOTCH2NLC* gene [5]. We excluded patients who were lost to follow-up. Patients were enrolled in August 2024. The detailed clinical data were obtained from the patients or relatives and medical records. Neurological examinations were performed by neurologists. Sex reported in this study is defined based on assigned at birth. Determinations of ethnicity was based on self-report. Spirometry and BDR testing were ordered by clinicians and conducted by medical technologists.

### Evaluations of the respiratory functions

The procedure of the evaluations is shown in Fig. 1. The patients with NIID diagnosed by histopathological studies and genetic tests were included in this study (Fig. 1, box 1). The clinicians asked the patients or relatives whether the patients had histories of asthma in childhood, histories of treatment for asthma, wheezing, persistent cough, or repetitive paroxysmal dyspnoea (Fig. 1, box 2). If the patients had even one of them, the clinicians additionally conducted the BDR testing. First of all, if possible, we measured the fractional concentration of exhaled nitric oxide (FeNO) using the commercially available analyser, NIOX VERO (Aerocrine AB, Solna, Sweden) or NObreath (BN, Bedfont, United Kingdom) (Fig. 1, box 3-1). We then performed the BDR testing consisting of 1st spirometry test, administration of the bronchodilator, and 2nd spirometry test, in order. In 1st spirometry, we measured VC, IC, FVC, FEV_1_, FEV_1_/ FVC, PEF, and MMF. Then, we calculated the ratio of the measured to predicted values of VC [18], FVC [18], FEV_1_ [19], FEV_1_/FVC [19], PEF [20], and MMF [21] (expressed as %VC, %FVC, %FEV_1_, %FEV_1_/FVC, %PEF, and %MMF, respectively) (Fig. 1, box 3-2). After 1st spirometry, the patients inhaled the beta 2 agonist (procaterol 20–30 µg or salbutamol 200 µg) (Fig. 1, box 3-3). Fifteen to 30 minutes later, we lastly conducted 2nd spirometry remeasuring or recalculating the abovementioned items examined in 1st spirometry (VC, IC, and %VC were checked, if possible.) (Fig. 1, box 3-4). We calculated the change amount (i.e. the value obtained by subtracting each value in 1st spirometry from that in 2nd spirometry) and change rate (i.e. the ratio of each change amount to the value in 1st spirometry). The evaluations were conducted from September 2024 to December 2024.

**Fig. 1.**
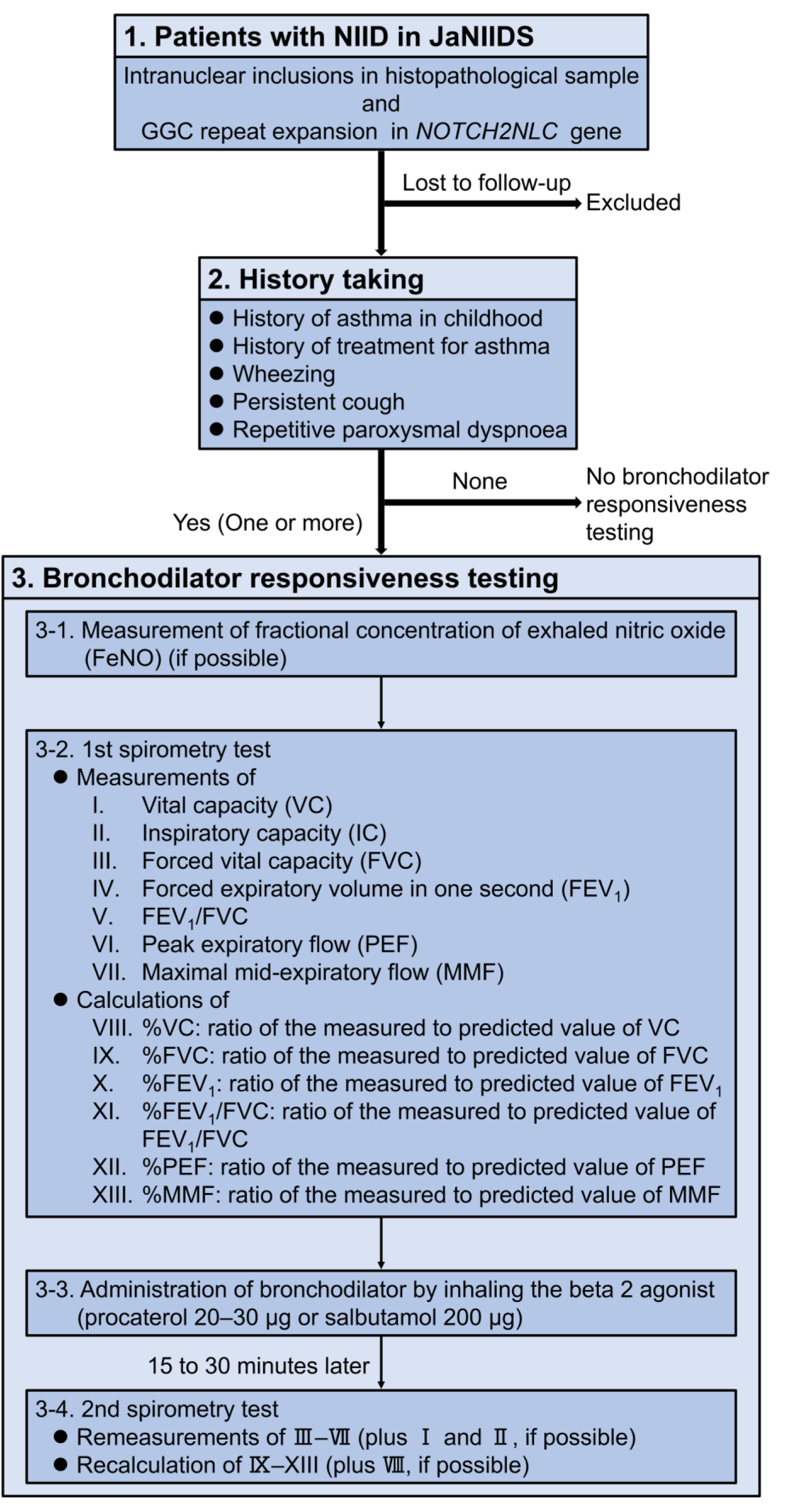
Patients and procedure for the bronchodilator responsiveness testing. We diagnosed patients as having neuronal intranuclear inclusion disease with an ante-mortem histopathological study and confirming GGC repeats expansion in *NOTCH2NCL* gene (box 1). We excluded patients who were lost to follow-up. The enrolled patients were asked some questions concerning respiratory symptoms (box 2). If they had one or more, we additionally conducted bronchodilator responsiveness testing (box 3). Before the 1st spirometry, we measured the fractional concentration of exhaled nitric oxide, if possible (box 3-1). Then we conducted the 1st spirometry test (box 3-2), administered the bronchodilator (box 3-3), and conducted the 2nd spirometry test (box 3-4), in order. JaNIIDS, the Japanese Consortium for neuronal intranuclear inclusion disease study; NIID, neuronal intranuclear inclusion disease.

### Statistical analysis

Data are expressed as number (%, 95% CI) or median (interquartile range). The Mann–Whitney U-test was used to compare continuous variables. We used the Wilcoxon signed-rank test for comparing the values in 1st and 2nd spirometry. Hypothesis tests were 2-sided. The significance level was set at *p* < 0.05. The statistical analyses were performed excluding missing data. The analyses were performed using EZR (Saitama Medical Center, Jichi Medical University, Saitama Japan), which is a graphical user interface for R (The R Foundation for Statistical Computing, Vienna, Austria, version 4.2.2) [21].

### Ethical approval and consent to participate/publish

Study protocols were approved by the Research Ethics Committee of Aichi Medical University (approval number: 2021-065 and 2021-175), and the study was performed in accordance with the standards of the 1964 Helsinki Declaration and its later amendments. Prior to the inclusion into this study, written informed consent was obtained from patients or their relatives according to the Declaration of Helsinki. Participants did not receive a stipend. Patients or their relatives signed informed consent regarding publishing their data.

## Results

Two hundred and thirty patients participated in JaNIIDS. We excluded 179 patients due to lacking of follow-up. Fifty-one patients were enrolled from the 29 hospitals in Japan (Fig. 2). Of the 51 patients, two had already died at the time of history taking, but we were able to obtain the medical histories from their relatives. Twenty (39.2%) were males, and 31 (60.8%) were females. All 51 patients were East-Asian. The median age at the enrolment was 71 (66–74) years. Of the 51 enrolled patients, 17 (33.3%, 95% CI 20.8% to 47.9%) patients had at least one history of asthma (Fig. 2, a). Clinical data and respiratory histories of the 17 patients (A–Q) are indicated in Table 1. Six (35.3%) were males, and 11(64.7%) were females. The median age was 64 (54–71) years (Patient P had already died at the time of history taking). Patients B, E, and G presented body mass index ≧ 25 kg/m^2^, the value classified as overweight or obesity [23] whereas patients C, J, and K presented that < 18.5 kg/m^2^, the value classified as underweight [23]. All 17 patients had GGC repeat expansion in the *NOTCH2NLC* gene, though the exact number of the GGC repeats were indeterminable in patients G and J. Twelve patients (A, B, E, G, H, I, J, K, L, O, P, and Q) had miosis. Three patients (B, D, and O) had some allergies, and two patients (O and P) had history of smoking. Two patients (J and N) had history of asthma in childhood and seven patients (B, F, J, K, M, N, and O) had history of treatment for asthma. Wheezing, persistent cough, and repetitive paroxysmal dyspnoea were complained of as current respiratory symptoms in 4 (F, J, M, and N), 15 (except F and O), and 3 (J, K, and M) patients, respectively (Table 1).

**Fig. 2.**
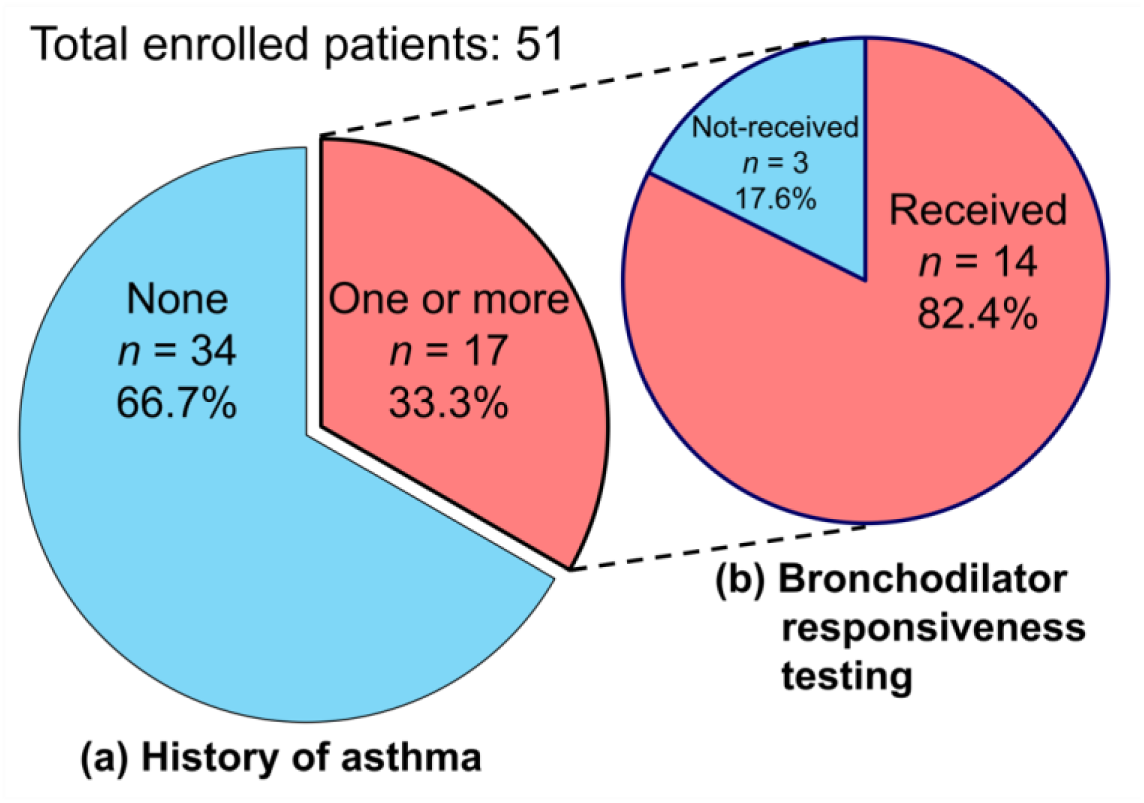
Outline of enrolled patients. Fifty-one patients were enrolled from the 29 hospitals in Japan. (a) Of the 51 patients, 17 (33.3%, 95%CI 20.8% to 47.9%) patients had one or more histories of asthma. (b) Of the 17 patients with one or more histories of asthma, 14 (82.4%) patients received bronchodilator responsiveness testing.

**Table 1.** Clinical findings of the patients with at least one respiratory symptom.

|  | A | B | C | D | E | F | G | H | I | J | K | L | M | N | O | P | Q |
| --- | --- | --- | --- | --- | --- | --- | --- | --- | --- | --- | --- | --- | --- | --- | --- | --- | --- |
| Sex (M, male; F, female.) | F | F | F | F | M | M | F | F | M | F | F | F | M | M | F | M | F |
| Body mass index (kg/m<sup>2</sup>) | 20.0 | 25.0 | 16.9 | 20.0 | 27.4 | 24.9 | 29.8 | 23.1 | 23.1 | 16.5 | 16.1 | 19.5 | 19.1 | 19.0 | NA | NA | NA |
| GGC repeats in *NOTCH2NLC* | 157 | 110 | 108 | 94 | 129 | 77 | ..<sup>a</sup> | 85 | 85 | ..<sup>a</sup> | 180 | 97 | 137 | 110 | 79 | 82 | 87 |
| Miosis | + | + | NA | - | + | NA | + | + | + | + | + | + | - | NA | + | + | + |
| Allergy | - | + | - | + | - | NA | - | - | - | - | NA | - | - | NA | + | NA | NA |
| Past or current smoking | - | - | NA | - | - | - | - | - | - | - | - | - | - | - | + | + | - |
| History taking |  |  |  |  |  |  |  |  |  |  |  |  |  |  |  |  |  |
| History of asthma in childhood | - | - | - | - | - | - | - | - | - | + | - | - | - | + | - | - | - |
| History of treatment for asthma | - | + | - | - | - | + | - | - | - | + | + | - | + | + | + | - | - |
| Wheezing | - | - | - | - | - | + | - | - | - | + | - | - | + | + | - | - | - |
| Persistent cough | + | + | + | + | + | - | + | + | + | + | + | + | + | + | - | + | + |
| Repetitive paroxysmal dyspnoea | - | - | - | - | - | - | - | - | - | + | + | - | + | - | - | - | - |

The first line (A–Q) indicates patients. ".." means "undeterminable". "+" or "-" indicates having the history/symptom or not, respectively. The patient P had already died before the evaluation. Patients O and Q could not perform the bronchodilator responsiveness testing due to severe cognitive impairments and a bedridden state, respectively. <sup>a</sup>Though the number of GGC repeats was undeterminable in patients G and J, the expansions themselves have been confirmed. *NA* not applicable.

Of the 17 patients with at least one respiratory symptom, the BDR testing was conducted in 14 patients (A–N in Table 1, 2, and 3, 82.4%) (Fig. 2, b). Patients O and Q could not perform the tests due to severe cognitive impairments and a bedridden state, respectively. Patient P had already died before the evaluation.

The results of FeNO and 1st spirometry in patients A–N are shown in Table 2. FeNO was measured in nine (B, C, F, G, H, I, K, L, and N) of 14 patients (Table 2). Median of FeNO was 15 (10–21) ppb. Most NIID patients in whom FeNO could be measured showed < 25 ppb, the normal value proposed in the American Thoracic Society guideline [24]. Only one patient (G) showed the indeterminant value of FeNO (34 ppb). No NIID patients showed > 50 ppb, the elevated value (Table 2). FEV_1_/FVC was < 70% in four patients (F, G, H, and N) (Table 2).

**Table 2.** FeNO and 1st spirometry test in the bronchodilator responsiveness testing.

|  | A | B | C | D | E | F | G | H | I | J | K | L | M | N |
| --- | --- | --- | --- | --- | --- | --- | --- | --- | --- | --- | --- | --- | --- | --- |
| FeNO (ppb) | NA | 8 | 21 | NA | NA | 10 | 34 | 22 | 8 | NA | 14 | 15 | NA | 21 |
| VC (L) | 2.33 | 3.14 | 3.24 | 2.21 | 3.41 | 1.57 | 2.85 | 2.08 | 2.63 | 2.32 | 2.55 | 2.07 | 3.35 | 2.68 |
| IC (L) | 1.29 | 2.85 | 2.07 | 1.90 | 2.22 | 1.37 | 2.28 | 1.45 | NA | 1.41 | 1.56 | 1.25 | 2.18 | 1.60 |
| FVC (L) | 2.32 | 2.91 | 3.30 | 2.34 | 3.49 | 1.47 | 2.88 | 2.09 | 2.57 | 2.37 | 2.43 | 2.03 | 3.56 | 2.68 |
| FEV<sub>1</sub> (L) | 1.97 | 2.35 | 2.64 | 1.97 | 2.98 | 0.79 | 1.98 | 1.42 | 1.83 | 1.92 | 2.09 | 1.55 | 3.12 | 1.73 |
| FEV<sub>1</sub>/FVC (%) | 84.91 | 80.76 | 80.00 | 84.19 | 85.39 | 53.74 | 68.75 | 67.94 | 71.20 | 81.01 | 86.01 | 76.35 | 87.64 | 64.55 |
| PEF (L/s) | 2.84 | 4.99 | 5.32 | 4.22 | 6.40 | 2.19 | 4.21 | 2.55 | 3.21 | 3.13 | 4.00 | 4.20 | 8.02 | 2.99 |
| MMF (L/s) | 1.88 | 2.17 | 2.17 | 2.27 | 2.73 | 0.49 | 1.06 | 0.83 | 1.36 | 1.77 | 2.46 | 1.15 | 3.98 | 0.96 |
| %VC (%)<sup>a</sup> | 88.26 | 129.75 | 113.29 | 99.55 | 97.43 | 51.48 | 125.55 | 98.11 | 78.27 | 90.98 | 85.57 | 85.89 | 88.16 | 73.63 |
| %FVC (%)<sup>a</sup> | 87.88 | 120.25 | 115.38 | 105.41 | 99.71 | 48.20 | 126.87 | 98.58 | 76.49 | 92.94 | 81.54 | 84.23 | 93.68 | 73.63 |
| %FEV<sub>1</sub> (%)<sup>a</sup> | 84.55 | 120.51 | 98.51 | 123.13 | 107.19 | 39.90 | 117.16 | 99.30 | 72.33 | 88.48 | 72.82 | 80.73 | 93.41 | 57.67 |
| %FEV<sub>1</sub>/FVC (%)<sup>a</sup> | 105.31 | 107.85 | 99.22 | 115.65 | 123.68 | 85.71 | 92.78 | 93.32 | 105.40 | 104.89 | 105.65 | 100.55 | 113.46 | 92.51 |
| %PEF (%)<sup>a</sup> | 36.79 | 70.88 | 66.83 | 63.94 | 67.02 | 26.42 | 62.00 | 39.29 | 34.78 | 42.24 | 49.38 | 58.91 | 77.79 | 30.54 |
| %MMF (%)<sup>a</sup> | 58.93 | 86.45 | 63.64 | 106.57 | 77.34 | 18.77 | 46.49 | 41.09 | 41.46 | 62.54 | 68.52 | 44.75 | 89.64 | 25.88 |

The first line (A–N) indicates patients. <sup>a</sup>The ratio of each measured to predicted value. *FeNO* fractional concentration of exhaled nitric oxide, *FEV<sub>1</sub>* forced expiratory volume in one second, *FVC* forced vital capacity, *IC* inspiratory capacity, *MMF* maximal mid-expiratory flow, *NA* not applicable, *PEF* peak expiratory flow, *VC* vital capacity.

The results of BDR testing are indicated in Table 3, Table 4, and Fig. 3. Table 3 shows the change amount and rate of FVC, FEV_1_, PEF, and MMF by the administration of the bronchodilator in each patient. FVC increased in eight patients (B, C, D, E, F, H, J, and M). Three patients (E, F, and M) presented an increase in FVC ≧ 0.20 L. Additionally, the change rate of FVC in patient F was over 12%. FEV_1_ increased in 13 patients (except L). Three patients (H, I, and N) presented an increase in FEV_1_ ≧ 0.20 L and 12%. The change rate of FEV_1_ in Patient F was over 12%. PEF increased in ten patients (A, B, D, G, H, I, J, L, M, and N), and three patients (A, H, and J) had a change rate over 20%. MMF increased in 12 patients (except D and F) (Table 3).

**Fig. 3.**
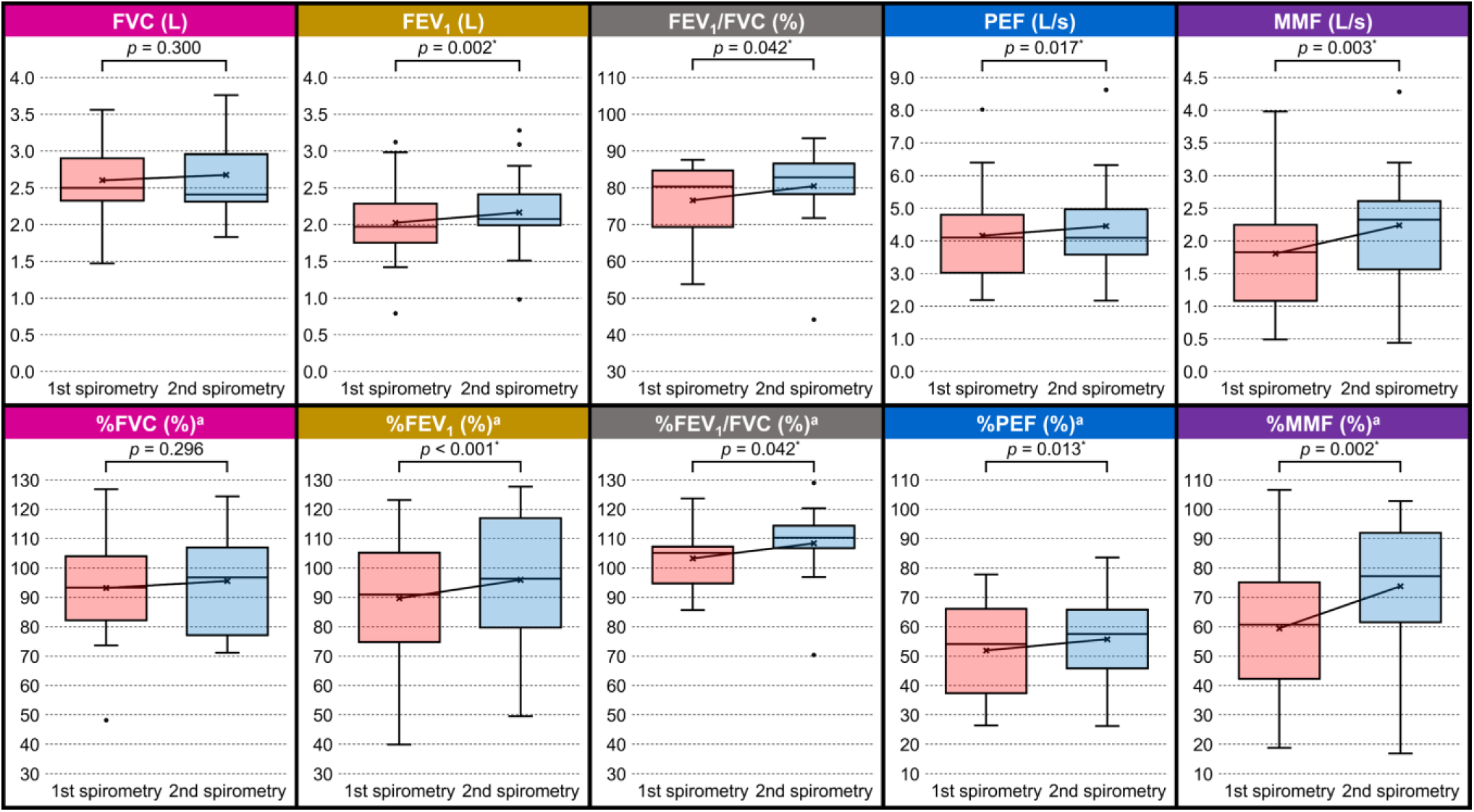
Boxplot of bronchodilator responsiveness testing result. A red box located in a left side in each box-plot shows the result of the 1st spirometry whereas a blue one in right shows the result of the 2nd spirometry. A cross mark inside each box shows the mean value. In each measured or calculated value, pre- and post-mean values (cross marks) are connected by a line across red and blue boxes. A horizontal line inside each box shows the median. Top and lower ends of each box indicate third and first quartiles, respectively. Upper and lower whiskers indicate maximum and minimum, respectively. Plots outside some boxes are outliers. A Wilcoxon signed-rank test was used for comparison of the data of 1st and the 2nd spirometry. ^a^The ratio of each measured to predicted value. \**p* < 0.05. FEV_1_, forced expiratory volume in one second; FVC, forced vital capacity; MMF, maximal mid-expiratory flow; PEF, peak expiratory flow.

**Table 3.** Change amount and rate in the bronchodilator responsiveness testing.

|  | A | B | C | D | E | F | G | H | I | J | K | L | M | N |
| --- | --- | --- | --- | --- | --- | --- | --- | --- | --- | --- | --- | --- | --- | --- |
| FVC |  |  |  |  |  |  |  |  |  |  |  |  |  |  |
| change amount (L)<sup>a</sup> | -0.03 | 0.10 | 0.14 | 0.04 | 0.23 | 0.75 | -0.08 | 0.11 | -0.16 | 0.04 | -0.02 | -0.20 | 0.20 | -0.09 |
| change rate (%)<sup>b</sup> | -1.29 | 3.44 | 4.24 | 1.71 | 6.59 | 51.02 | -2.78 | 5.26 | -6.23 | 1.69 | -0.82 | -9.85 | 5.62 | -3.36 |
| FEV<sub>1</sub> |  |  |  |  |  |  |  |  |  |  |  |  |  |  |
| change amount (L)<sup>a</sup> | 0.17 | 0.14 | 0.16 | 0.02 | 0.11 | 0.19 | 0.03 | 0.28 | 0.27 | 0.13 | 0.09 | -0.04 | 0.16 | 0.26 |
| change rate (%)<sup>b</sup> | 8.63 | 5.96 | 6.06 | 1.02 | 3.69 | 24.05 | 1.52 | 19.72 | 14.75 | 6.77 | 4.31 | -2.58 | 5.13 | 15.03 |
| PEF |  |  |  |  |  |  |  |  |  |  |  |  |  |  |
| change amount (L/s)<sup>a</sup> | 0.71 | 0.40 | -0.17 | 0.22 | -0.08 | -0.02 | 0.16 | 0.87 | 0.46 | 0.69 | -0.29 | 0.25 | 0.60 | 0.27 |
| change rate (%)<sup>b</sup> | 25.00 | 8.02 | -3.20 | 5.21 | -1.25 | -0.91 | 3.80 | 34.12 | 14.33 | 22.04 | -7.25 | 5.95 | 7.48 | 9.03 |
| MMF |  |  |  |  |  |  |  |  |  |  |  |  |  |  |
| change amount (L/s)<sup>a</sup> | 0.74 | 0.41 | 0.28 | -0.17 | 0.47 | -0.05 | 0.17 | 0.72 | 1.67 | 0.43 | 0.09 | 0.35 | 0.30 | 0.65 |
| change rate (%)<sup>b</sup> | 39.36 | 18.89 | 12.90 | -7.49 | 17.22 | -10.20 | 16.04 | 86.75 | 122.79 | 24.29 | 3.66 | 30.43 | 7.54 | 67.71 |

The first line (A–N) indicates patients. <sup>a</sup>The values in 2nd - those in 1st spirometry. <sup>b</sup>The change amount / the value of 1st spirometry. *FEV<sub>1</sub>* forced expiratory volume in one second, *FVC* forced vital capacity, *MMF* maximal mid-expiratory flow, *PEF* peak expiratory flow.

**Table 4.**
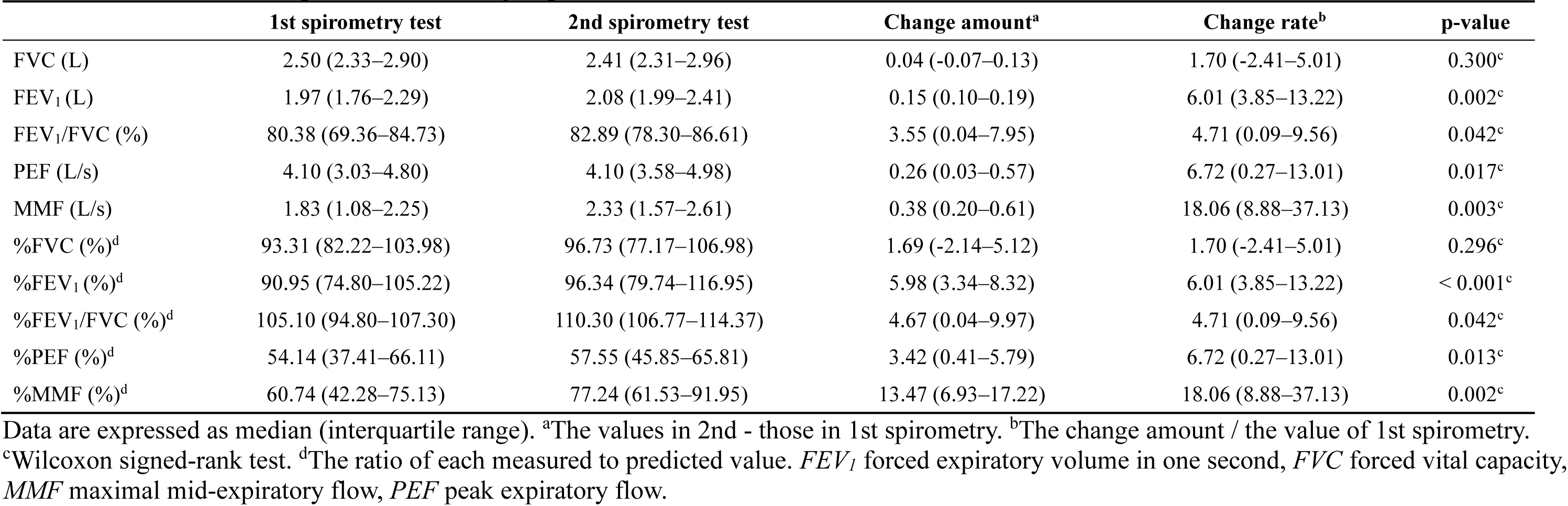
Bronchodilator responsiveness testing in patients A–N.

|  | 1st spirometry test | 2nd spirometry test | Change amount<sup>a</sup> | Change rate<sup>b</sup> | p-value |
| --- | --- | --- | --- | --- | --- |
| FVC (L) | 2.50 (2.33–2.90) | 2.41 (2.31–2.96) | 0.04 (-0.07–0.13) | 1.70 (-2.41–5.01) | 0.300<sup>c</sup> |
| FEV<sub>1</sub> (L) | 1.97 (1.76–2.29) | 2.08 (1.99–2.41) | 0.15 (0.10–0.19) | 6.01 (3.85–13.22) | 0.002<sup>c</sup> |
| FEV<sub>1</sub>/FVC (%) | 80.38 (69.36–84.73) | 82.89 (78.30–86.61) | 3.55 (0.04–7.95) | 4.71 (0.09–9.56) | 0.042<sup>c</sup> |
| PEF (L/s) | 4.10 (3.03–4.80) | 4.10 (3.58–4.98) | 0.26 (0.03–0.57) | 6.72 (0.27–13.01) | 0.017<sup>c</sup> |
| MMF (L/s) | 1.83 (1.08–2.25) | 2.33 (1.57–2.61) | 0.38 (0.20–0.61) | 18.06 (8.88–37.13) | 0.003<sup>c</sup> |
| %FVC (%)<sup>d</sup> | 93.31 (82.22–103.98) | 96.73 (77.17–106.98) | 1.69 (-2.14–5.12) | 1.70 (-2.41–5.01) | 0.296<sup>c</sup> |
| %FEV<sub>1</sub> (%)<sup>d</sup> | 90.95 (74.80–105.22) | 96.34 (79.74–116.95) | 5.98 (3.34–8.32) | 6.01 (3.85–13.22) | < 0.001<sup>c</sup> |
| %FEV<sub>1</sub>/FVC (%)<sup>d</sup> | 105.10 (94.80–107.30) | 110.30 (106.77–114.37) | 4.67 (0.04–9.97) | 4.71 (0.09–9.56) | 0.042<sup>c</sup> |
| %PEF (%)<sup>d</sup> | 54.14 (37.41–66.11) | 57.55 (45.85–65.81) | 3.42 (0.41–5.79) | 6.72 (0.27–13.01) | 0.013<sup>c</sup> |
| %MMF (%)<sup>d</sup> | 60.74 (42.28–75.13) | 77.24 (61.53–91.95) | 13.47 (6.93–17.22) | 18.06 (8.88–37.13) | 0.002<sup>c</sup> |

Data are expressed as median (interquartile range). <sup>a</sup>The values in 2nd - those in 1st spirometry. <sup>b</sup>The change amount / the value of 1st spirometry.
<sup>c</sup>Wilcoxon signed-rank test. <sup>d</sup>The ratio of each measured to predicted value. *FEV<sub>1</sub>* forced expiratory volume in one second, *FVC* forced vital capacity, *MMF* maximal mid-expiratory flow, *PEF* peak expiratory flow.

Table 4 and Fig. 3 indicate the statistical analyses for the BDR testing. After inhaling the beta 2 agonist, FEV_1_, FEV_1_/FVC, PEF, and MMF significantly increased by 0.15 L (6.01%, *p* = 0.002), 3.55% (4.71%, *p* = 0.042), 0.26 L/s (6.72%, *p* = 0.017), and 0.38 L/s (18.06%, *p* = 0.003), respectively. Furthermore, %FEV_1_, %FEV_1_/FVC, %PEF, and %MMF also significantly increased (*p* < 0.001, *p* = 0.042, 0.013, and 0.002, respectively) (Table 4 and Fig. 3). The median of the change rate in MMF (18.06 (8.88–37.13) %) was more than that in FEV_1_ (6.01 (3.85–13.22) %) (*p* = 0.044, Mann–Whitney U-test). The medians of %MMF in 1st spirometry (60.74 (42.28–75.13) %) was lower than that of %FEV_1_ (90.95 (74.80–105.22) %) (*p* = 0.027, Mann–Whitney U-test).

## Discussion

We comprehensively investigated the pulmonary conditions in patients with NIID by spirometry and BDR testing. After the administration of the bronchodilator, FEV_1_, FEV_1_/FVC, PEF, and MMF significantly increased. In BDR testing, FVC and FEV_1_ are primarily evaluated to determine the degree of improvement of airflow by the administration of the bronchodilator [15]. For the diagnosis of asthma, it is necessary to confirm airflow reversibility, with criteria in adults requiring an increase from baseline in FVC or FEV_1_ of ≧ 12% and ≧ 200 mL [25]. But even asthmatic patients do not always present ΔFEV_1_ > 12%- and 200 mL- changes [26]. In the present study, medians of change rate and amount of FEV_1_ (6.01% and 150 mL, respectively) did not satisfy the criteria; however, FEV_1_ significantly increased after the administration of the bronchodilator (*p* = 0.002) (Table 4 and Fig. 3). Furthermore, FVC and/or FEV_1_ increased in all patients expect one patient (L), and four patients (F, H, I, and N) satisfied the criteria of asthma (Table 3). We think NIID patients have asthma-like conditions. Though its reliability for the diagnosis of asthma is inferior to the spirometry, the diagnosis based on the increase in PEF ≧ 20% is acceptable in the criteria [25]. The present study showed a significant but less than criteria increase in PEF (6.72%, *p* = 0.017) as well as in FEV_1_ (Table 4 and Fig. 3). This result also supports the asthma-like conditions in NIID patients.

MMF reflects small airway function whereas FEV_1_ is used for an assessment of larger airways [16]. Reported mean %MMF in asthmatic patients is 49.9% [27] to 69% [28] whereas that in controls is 88.9% [27] to 93% [28], which is significantly lower in asthma [27, 28]. In addition, %MMF was lower than %FEV_1_ in asthma [29]. Moreover, MMF generally more improves than FEV_1_ in BDR testing especially in milder asthma [16]. In this study, the median of %MMF in the 1st spirometry (60.74%) was significantly lower than %FEV_1_ (90.95%) (*p* = 0.027). This results also support that NIID patients have asthma-like conditions. Furthermore, the change rate in MMF was more than in FEV_1_ (*p* = 0.044). We consider that the severity of asthma-like conditions in NIID is mild.

The symptoms of asthma include cough, wheezing, and shortness of breath, and are characterized by variability [25]. Asthma is heterogeneous, and some phenotypes are reported [25]. Cough-variant asthma predominantly presents paroxysmal coughs without other asthmatic symptoms such as wheezing and dyspnoea [30]. The variant also shows the variable expiratory air flow limitation and the therapeutic responsiveness to the inhaled corticosteroid and bronchodilators [30]. In our study, 13 of 14 patients who received the BDR testing had persistent coughs, and nine of 14 patients had only coughs as a present symptom (Table 1). Spirometry in cough-variant asthma is often normal [30], and six of the nine patients in our study had normal pulmonary function. Asthma-like conditions in NIID may be close to cough-variant asthma.

COPD is another obstructive pulmonary disorder presenting persistent respiratory symptoms including dyspnoea, cough, and wheezing [31]. In the BDR testing, the presence of a post-bronchodilator FEV_1_/FVC < 0.7 is necessary to diagnose COPD [31]. Most of the patients did not satisfy this criterion, and the median of the FEV_1_/FVC after the administration of bronchodilator was 0.8289 in the current study (Table 4). NIID patients seemed to have respiratory conditions more resembling asthma than COPD.

The asthma-like conditions of NIID may be resulted from the autonomic dysfunction caused by NIID. The efferent autonomic neurons regulate the calibres of the airways. The airways are contracted mainly by acetylcholine released from the parasympathetic postganglionic cholinergic neurons whereas the sympathetic neurons have bronchodilator functions [32]. In addition, parasympathetic components also have bronchodilator functions via transmitters such as nitric oxide and vasoactive intestinal peptide [32]. An increased cholinergic activity is known to be associated with asthma and COPD [32]. Various autonomic symptoms (e.g. miosis and bladder dysfunction) are reported in NIID patients [2]. In the autopsies of two NIID patients, neuronal loss and astrogliosis were observed in sympathetic ganglion neurons whereas not in dorsal vagal nucleus [8]. Additionally, in another autopsied case with bilateral miosis, almost normal findings of the Edinger-Westphal nucleus, which supplies parasympathetic fibres to eyes, were observed, and many intranuclear inclusions as well as neuronal loss were observed in the sympathetic ganglia [7]. The authors guess parasympathetic dominancy resulted in miosis [7]. In this study, many patients had miosis (Table 1). In NIID, sympathetic systems would be predominantly disturbed compared to parasympathetic. The sympathetic hypoactivity and relative parasympathetic hyperactivity may cause asthma-like symptoms.

In recent years, cerebellar ataxia with neuropathy and vestibular areflexia syndrome (CANVAS) has been reported as a neurodegenerative disease with respiratory symptoms accompanied by chronic cough [33–36]. CANVAS primarily presents with cerebellar ataxia, loss of vestibular reflexes, and peripheral neuropathy, and also been reported to be accompanied by chronic cough [37]. CANVAS patients exhibit pins and needles and have a hypersensitive reaction to the inhaled capsaicin challenge test [33], leading some reports to infer that hyperalgesia due to peripheral neuropathy is the cause of the chronic cough. On the other hand, pins and needles are rarely a problem in NIID [2, 8], and rather, peripheral neuropathy caused by damage to large myelinated fibres, such as loss of tendon reflexes and reduced vibration sense, is frequently exhibited [8]. Therefore, it is speculated that the underlying pathology of cough in NIID is different from that of CANVAS.

Asthma is usually characterized by chronic airway inflammations [25]. FeNO correlates with Type 2 airway inflammation, and its measurement is one of non-invasive ways to guess the presence of airway inflammation [24]. Cut points for FeNO in adults is set as follows: low FeNO < 25 ppb (Eosinophilic airway inflammation is unlikely.) and high FeNO > 50 ppb (Eosinophilic airway inflammation is likely.). FeNO between 25 and 50 ppb should be interpreted cautiously considering the clinical context [38]. Some factors such as obesity and smoking lower FeNO value [24]. Of nine patients measured FeNO in our study, two had body mass index ≧ 25 kg/m^2^, and none had histories of smoking (Table 1 and Table 2). These factors may influence the FeNO values; however, the values were generally low, and in only one patient the FeNO exceeded 25 ppb but lower than 50 ppb (Table 2). The respiratory conditions in NIID may categorize in a subtype of non-Type 2 or Type 2-“low” asthma that an endotype without markers of Type 2 inflammation, such as eosinophilia [39]. We believe that airway inflammation is not strongly associated with the pathogenesis of asthma-like conditions in NIID patients, and the effect of steroids is thought to be limited. We speculate that one of the reasons why NIID patients in our cohort discontinued asthma treatment in the past may be that they did not feel the effects of the steroid-based asthma treatment.

The prevalence of asthma symptoms among 20–79-year-old in Japan have been reported as up to 12.5% [40]. Compared to this report, the prevalence of respiratory symptoms in NIID patients observed in this study (33.3%, 95% CI 20.8% to 47.9%) is high, and treatments are necessary for improvements in their quality of life. According to international guidelines on asthma treatment, medicines for asthma include inhaled corticosteroid, short or long-acting beta 2 agonists, long-acting muscarinic antagonists, and biologics (e.g. anti-IgE and anti-interleukin 5), and inhaled corticosteroid-containing therapies are recommended from the time of diagnosis [25]. However, we think corticosteroids with anti-inflammatory effects and biologics acting on the immune system may be less effective in NIID patients. We guess the relative parasympathetic nervous dominant state results in asthma-like conditions in NIID. Thus, beta 2 agonists and muscarinic antagonists may be more effective than other agents. As to the acting times of the drugs, it is reasonable to use long-acting agents (e.g. salmeterol and tiotropium [25]) for the control of respiratory symptoms in NIID patients because NIID is a chronic progressive disease without disease-modifying drugs, and autonomic imbalance seemed to be permanent.

This study has some limitations. Firstly, some nervous symptoms caused by NIID may influence results of the spirometry. It may be difficult to understand the methods well in patients with dementia, and to conduct the sufficient forced expiration in those with muscle weakness. Secondly, some items such as FeNO and IC were not available in some patients. Thirdly, biomarkers of Type 2 inflammation other than FeNO (e.g. peripheral blood eosinophils and serum total and allergen-specific IgE [24]) and chest CT scan were not evaluated in this study. Lastly, BDR testing was conducted only in the NIID patients with respiratory symptoms, thus we lacked control data in NIID patients without respiratory symptoms. However, requiring individuals without symptoms to inhale the beta 2 agonist poses an ethical issue.

Despite these limitations, this study has the strength of presenting detailed comprehensive data concerning respiratory conditions and BDR in NIID patients. We propose that clinicians consider the possibility of NIID if patients have asthmatic symptoms (especially persistent cough without responsiveness to corticosteroid) with chronic neurological symptoms including dementia and neuropathy. In future research, it is necessary to investigate actual effects of beta 2 agonists or muscarinic antagonists for clinical implementation. In addition, further studies, such as pathological investigations of thoracic sympathetic ganglia and parasympathetic systems, are also necessary to elucidate the mechanism of asthma-like conditions in NIID patients.

## Conclusions

Patients with NIID have mild asthma-like conditions. This condition may be not due to chronic airway inflammation but due to sympathetic nervous dysfunction caused by NIID. Immunomodulators such as corticosteroid may not improve respiratory symptoms in NIID whereas beta 2 agonists or muscarinic antagonists may improve those symptoms.

## Data availability

The data of this study are not publicly available due to the necessity to obtain the approval of The Research Ethics Committee of Aichi Medical University. The data are available from the corresponding author after the approval of The Research Ethics Committee of Aichi Medical University, upon reasonable request.

## Abbreviations

BDR: bronchodilator responsiveness
CANVAS: cerebellar ataxia with neuropathy and vestibular areflexia syndrome
COPD: chronic obstructive pulmonary disease
FeNO: fractional concentration of exhaled nitric oxide
FEV_1_: forced expiratory volume in one second
FVC: forced vital capacity
IC: inspiratory capacity
JaNIIDS: the Japanese Consortium for neuronal intranuclear inclusion disease study
MMF: maximal mild-expiratory flow
NIID: neuronal intranuclear inclusion disease
PEF: peak expiratory flow
VC: vital capacity

## Acknowledgements

We gratefully thank members of JaNIIDS group for data collection: Arisa Yamamoto (NHO Asahikawa Medical Center), Takayuki Katayama (Asahikawa City Hospital), Mai Fujikura (SAISEIKAI OTARU Hospital), Atsushi Sakurai (Isesaki Municipal Hospital), Toshiyuki Hayashi (Nippon Medical School), Manabu Araki (Kawakita General Hospital), Keizo Sugaya (Tokyo Metropolitan Neurological Hospital), Takamura Nagasaka (Kofu neurosurgical hospital), Yuzo Tanaka (NHO Shizuoka Medical Center), Takanobu Kita (Ichinomiyanishi Hospital), Nobuyuki Iwade (Yokkaichi Municipal Hospital), Takao Kiriyama (Nara Medical University), Yosuke Osakada (Okayama University Graduate School of Medicine, Dentistry and Pharmaceutical Sciences), Megumi Toko (Hiroshima University Hospital), Motoharu Kawai (Neuromuscular Center Yoshimizu Hospital), Yoshiaki Takahashi (Kagawa Prefectural Central Hospital), Tomoyasu Matsubara (Tokushima University Graduate School of Biomedical Science), Noriyuki Miyaue (Ehime University Graduate School of Medicine), Masahiro Shijo (Kyushu Central Hospital Of The Mutual Aid Association Of Public School Teachers), Toshihiro Ide (Saga university), Masaaki Yoshikawa (Saga university), and Hidenori Matsuo (NHO Nagasaki National Hospital). We also gratefully thank Hiroshi Takashima (Kagoshima University Graduate School of Medical and Dental Sciences) and Satoko Kumada (Tokyo Metropolitan Neurological Hospital) for supports of this study. We are grateful to the patients with NIID, their families, and everyone involved in in this study and JaNIIDS. And the authors gratefully thank institute of comprehensive medical research, division of advanced research promotion, Aichi Medical University for the *NOTCH2NLC* GGC repeat analysis.

## Author contributions

DT and NT acquired and curated data, analysed and interpreted the results, and wrote the paper. JS and SI conceived and designed this study, and analysed and interpreted the data. JS managed the study, acquired funding, examined NIID patients, performed the skin biopsies and genetic analyses, constructed the NIID cohort, recruited patients, and analysed and interpreted the data. CT performed skin histological analysis, genetic testing according to NIID diagnosis. AA, YR, TF, KH, HI, TakashiKa, TosM, FT, and YaI interpreted the data and reviewed the paper. MasA and YH performed genetic testing according to NIID diagnosis, interpreted the data, and review the paper. RK, HO, and GS reviewed the paper. HirM analysed the data. MiK, JI, HY, TakashiKu, AtsuhS, AN, KN, KaS, TF, KH, HI, HK, TakashiKa, TosM, FT, and YuI history taking and/or BDR testing. All authors read and approved the final manuscript. All authors had full access to all the data in the study and had final responsibility for the decision to submit for publication.

